# A strategy of eculizumab treatment withdrawal with disease monitoring is cost-effective compared to lifelong treatment for eligible atypical haemolytic uraemic syndrome patients: an economic evaluation of the SETS aHUS trial

**DOI:** 10.1101/2025.07.14.25331491

**Authors:** Giovany Orozco-Leal, Luke Vale, Yemi Oluboyede, Andy Bryant, Chris Weetman, Ciara Kennedy, David Kavanagh, Edwin Wong, Jan Lecouturier, Len Woodward, Victoria Brocklebank, Sally Johnson, Sarah Dunn, Tom Chadwick, Neil S Sheerin

**Author notes:** **Correspondence to:** Giovany Orozco-Leal.

## Abstract

**Background and hypothesis:** Atypical haemolytic uraemic syndrome (aHUS) is a rare condition caused by compliment dysregulation. Eculizumab is an effective treatment for patients with aHUS, yet a lifelong treatment strategy is costly for health services and is of uncertain additional benefit to patients. This economic evaluation was conducted as part of stopping eculizumab treatment safely in aHUS (SETS aHUS) trial and assessed the cost-effectiveness of a lifelong delivery of eculizumab strategy compared with stopping treatment with a disease monitoring strategy over the long-term.

**Methods:** A Markov model was used to estimate cost and quality adjusted life years (QALYs) for the two strategies compared. The main source of data used was SETS aHUS, for quality-of-life, resource use estimates, and treatment probabilities. Time to treatment restart over an estimated patient lifetime was extrapolated from trial data using parametric survival functions.

**Results:** The eculizumab withdrawal and disease monitoring strategy changed QALYs by 0.22 (95% CrI: −0.7 to 1.25), and reduced costs per patient by £4 188 361 (95% CrI: -£6 390 713 to -£675 511) compared with the lifelong delivery of eculizumab. Survival was similar, with withdrawal patients presenting 0.0005 LYs less on average (95% CrI: −0.003 to 0) over an 80-year time horizon. The likelihood of withdrawal being more effective and less costly was 71%. Results were robust across multiple scenarios exploring uncertainties.

**Conclusion:** Treatment withdrawal with disease monitoring strategy is cost-effective compared with lifelong treatment with eculizumab. Its adoption is expected to substantially reduce costs per patient and may improve patient quality of life on average.

## Introduction

Atypical Haemolytic Uraemic Syndrome (aHUS) is a rare, life-threatening disease characterised by a severe inflammation of blood vessels and the formation of blood clots throughout the body leading to tissue injury and, subsequently, thrombotic microangiopathy (TMA). The disease more often manifests clinically by TMA involving the kidneys, and if left untreated can lead to kidney failure [1, 2].

In the UK, the incidence of aHUS is 0.4 to 0.5 cases per million per year. In 70% of these cases the disease is associated with an excessive activation of the complement system due to a genetic or acquired abnormality [2]. Eculizumab is part of a group of drugs known as C5 inhibitors; C5 being an important protein involved in complement activation. Thus, eculizumab prevents tissue damage from the excessive activation of the complement system [2].

In 2015, the National Institute for Health and Care Excellence (NICE) recommended eculizumab for the treatment of aHUS [2]. NICE recommended that treatment be lifelong but also recommended research to investigate the safety of dose adjustments and treatment discontinuation [2]. The introduction of eculizumab changed the treatment pathway and improved the prognosis of aHUS patients but treatment duration is still debated, with a dearth of studies investigating the optimal duration of treatment [3].

Treatment with eculizumab is not without risk as its delivery is associated with an increased risk of meningococcal infection [4]. The most salient issue stimulating the debate on treatment duration with eculizumab is however its prohibitive price. Lifelong treatment duration comes with a substantial cost to health services and patients, even in countries where treatment is free at the point of use due to the frequency of hospital appointments that patients must attend [3, 5].

The Stopping Eculizumab Treatment Safely in aHUS (SETS aHUS) trial came about as a response to NICE’s call for a study assessing the safety of stopping eculizumab treatment [1, 2]. SETS aHUS was a stage 2, single-arm, non-blinded assessment of the safety and cost-effectiveness of eculizumab withdrawal in patients with aHUS using a Bayes factor single-arm design [6]. Due to the trial methodology and the small sample size (n =39), a within-trial economic evaluation was not deemed appropriate. Instead, the trial collected costs and health outcomes for the trial sample and for a comparator group made up of patients under treatment maintenance. The purpose of this data collection was to provide data to assess the cost-effectiveness of eculizumab withdrawal with a disease monitoring protocol over the long term.

This paper reports how we used clinical and health economic data from the SETS aHUS trial along with data from the literature to construct an economic model and assessed the cost-effectiveness of withdrawal from eculizumab, substituted by a system of protocolised surveillance and a strategy for treatment reintroduction, compared with the lifelong delivery of eculizumab.

## Materials and Methods

Full details on the methodology used for the economic analysis are in the health economic analysis plan (Appendix 1). Further details of the trial protocol are reported in Dunn *et al.,* 2022 [1], and its primary results in Bryant *et al.,* 2024 [7].

### Population

The target population was adult patients diagnosed with aHUS receiving eculizumab to treat the disease in native kidneys with either normal renal function (CKD stage 0) or chronic kidney disease stages 1-3 (CKD 1-3). Patients undergoing transplant, or patients at or below CKD stage 4 were not included. In the base-case analysis a cohort of patients aged 20 years old; comprising 51% males/49% females was modelled based on the trial sample and treatment maintenance cohort of SETS aHUS [7].

### Setting and location

The setting was the UK national health service (NHS). After eculizumab initiation, patients maintaining treatment were eligible to have it delivered at home or at the hospital by a specialist nurse. Patients under treatment withdrawal attended inpatient visits to the hospital to assess their disease status. The restart of treatment was also delivered in the inpatient setting.

### Intervention and comparator

Following the trial protocol, patients in all arms of the model had completed at least 6 months of eculizumab treatment, including vaccination against meningococcal infections and other antibiotic prophylaxis [1]. At the start of the model, patients in the withdrawal arm ceased eculizumab treatment and commenced disease monitoring (consistent with the SETS aHUS trial protocol [1]), for the rest of their life or until treatment was restarted. If treatment was restarted it was maintained for the rest of the patient lifetime. Patients in the control arm of the model remained on treatment with eculizumab for the rest of their life.

### Model structure

To inform the model structure, a literature review was conducted in June 2023 using EMBASE and Web of Science to identify model-based economic evaluations on aHUS. Three economic evaluations were identified [8–10], of which two were model-based analyses [8, 9] in addition to technology appraisals conducted by NICE (HST1 and TA710) [11, 12].

The *de novo* economic model was built in Microsoft Excel V2016 (Figure 1). A Markov model structure based on the most recent submission to NICE was considered the best approach [11], informed by data from the SETS aHUS trial. Health states in the model represent CKD stages based on eGFR scores, from normal kidney function (eGFR of 90 or above, CKD stage 1) to end-stage renal disease (ESRD, kidney failure with eGFR less than 15) [13]. The first health state grouped patients with CKD stages 0 to 2, the next health states were CKD 3, CKD 4, ESRD, kidney transplant, and a post-transplant state. Patients entered the model at either CKD 0-2 or CKD 3, based on the disease distribution in the SETS aHUS trial. CKD 0-2 patients could stay off treatment, restart treatment, progress to CKD 3 and restart treatment, or move to the death state. CKD 3 patients could stay off treatment, restart treatment at CKD 3 disease, progress to CKD 4 and restart treatment, or move to the death state. Patients at CKD 0-2 and CKD 3 receiving treatment were assumed to stay in their CKD health state for the rest of the time horizon.

**Figure 1.**
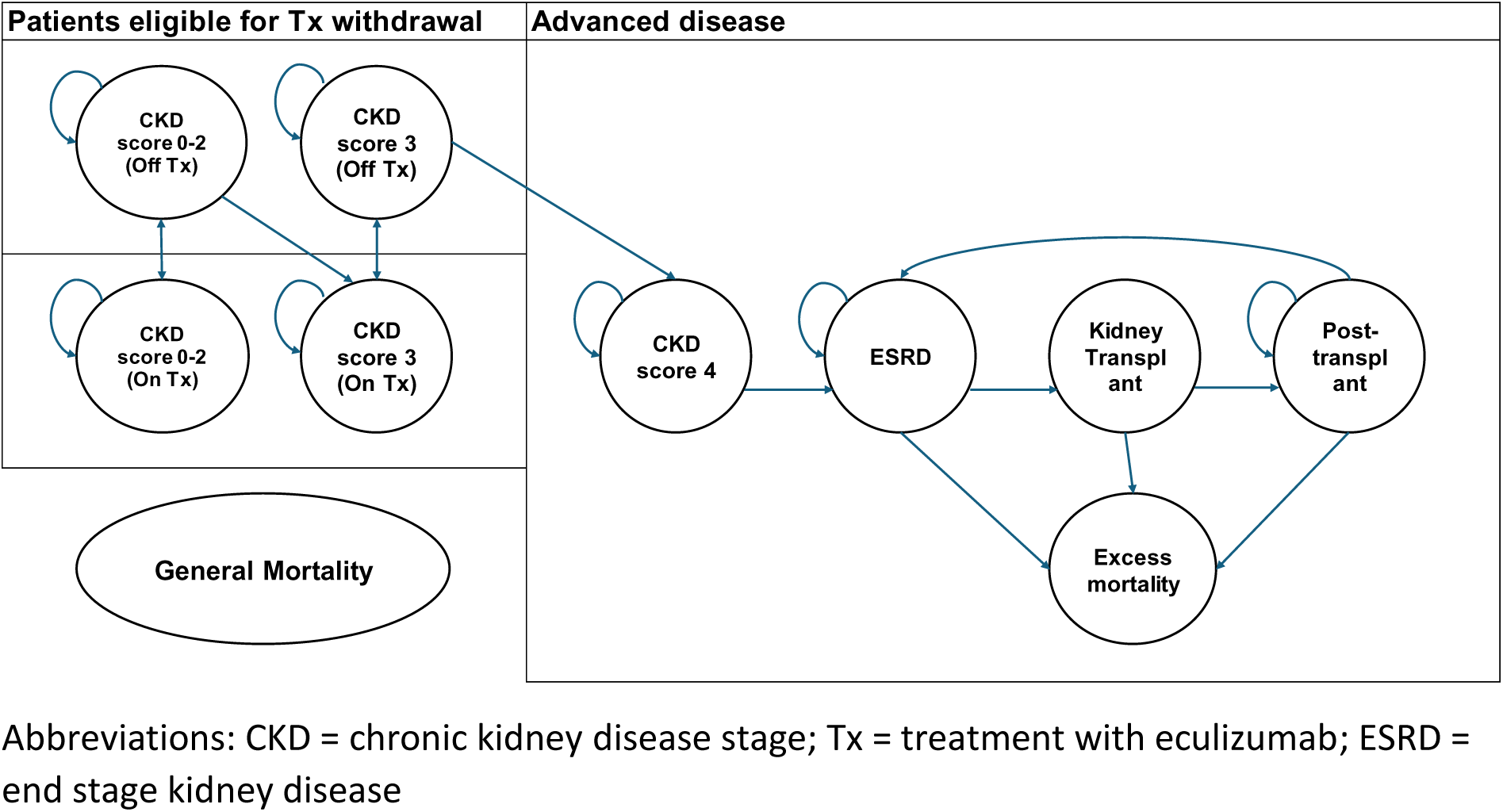
Model structure.

All patients progressing to CKD stage 4 or higher were assumed to continue receiving eculizumab treatment and could stay in CKD 4 until death or disease progression to ESRD. When patients progressed to ESRD they could stay in ESRD, or receive a kidney transplant, which is a tunnel state to the post-transplant state. Patients progressing to ESRD or beyond faced a higher risk of mortality due to disease severity and transplant-related causes. The model structure is presented in Figure 1.

A cycle length of 2 weeks and a half-cycle correction was used to capture treatment cycles and disease stages. A lifetime horizon was considered appropriate for the base-case analysis, ending the simulation when surviving patients were 100 years old.

### Health state transition probabilities

Disease distribution at baseline based on CKD stages was 96% (n=27) at CKD 0-2, and 4% at CKD 3 (n=1) corresponding to the baseline distribution in SETS aHUS. Time to event data from the trial showed 4 patients (14%) relapsed and restarted treatment after 2 years of eculizumab withdrawal, of which 1 patient had a primary outcome event with a persistent >20% fall in eGFR, while the rest recovered their kidney function after restarting treatment [7]. The probability of disease relapse over time was derived from fitting standard parametric functions to the Kaplan Meier data [14]. Due to the small sample size, the functions fitted included: exponential, Weibull, Gompertz, Log logistic, and lognormal [14]. The risk of disease progression from aHUS relapse was assumed constant over time. This is potentially a conservative assumption, since further data from the aHUS SETS trial cohort showed that the patient who experience a primary outcome event recovered after the study follow-up period [7].

The differences in Bayesian information criterion (BIC) scores, used to measure the goodness of fit to the data, were small across the five functions. The exponential function was judged as having the best BIC score; however, the Log-normal function was selected for the base-case due to its decreasing hazard over time in line with evidence from Nikolaidis G *et al* (2020) [15], moreover, its long-term predictions were a mid-point between the predictions considered (Figure 2).

**Figure 2.**
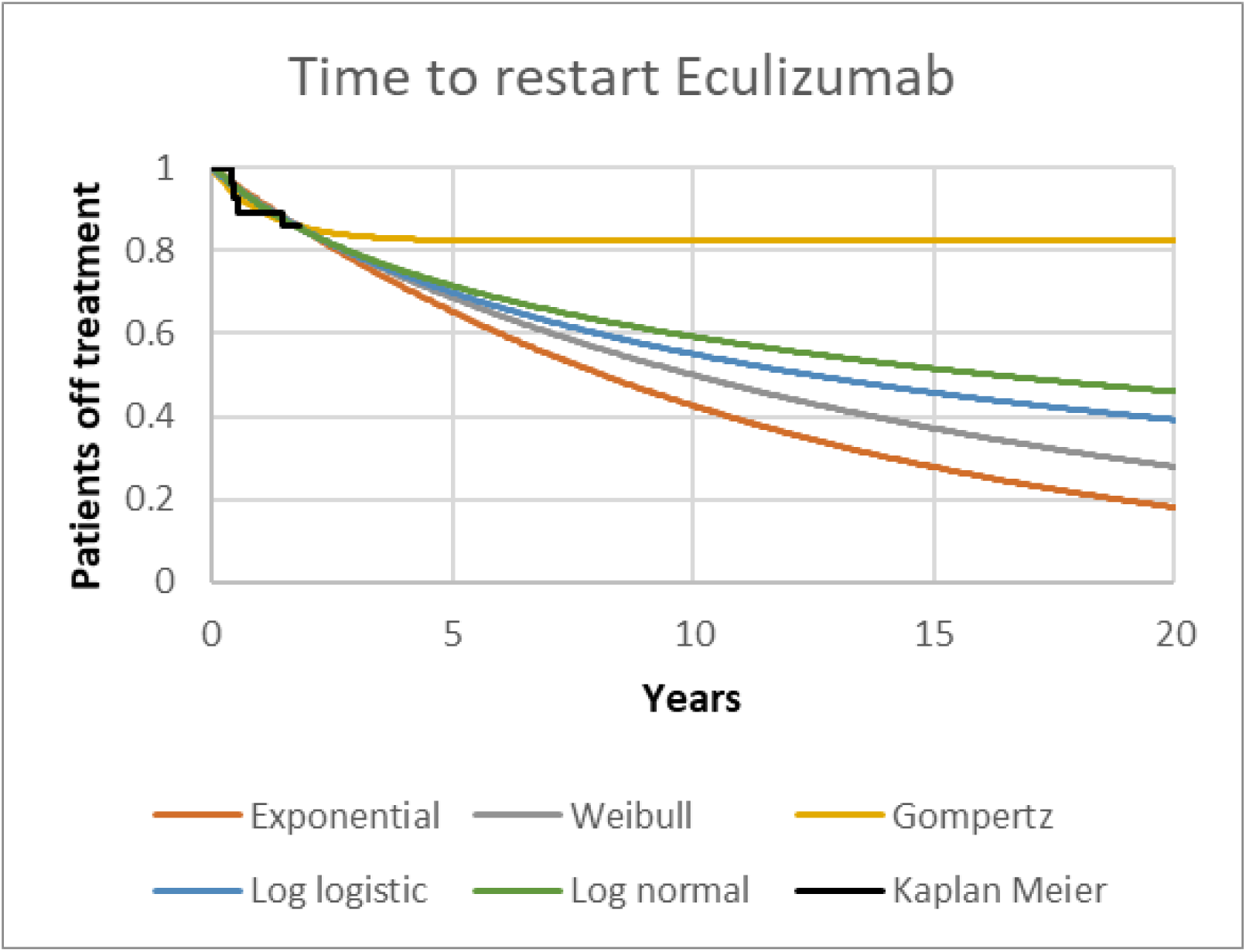

Patients at CKD 0-2 and CKD 3 who are receiving or restarting treatment with eculizumab are assumed to have no risk of progression. For patients at CKD stage 4, the risk of progressing to ESRD was based on the results from Rondeau *et al*., 2020 [16].

Patients face a mortality risk sourced from ONS life tables for the general UK population. Patients progressing to the health states of end stage renal disease (ESRD), transplant, and post-transplant care face an additional 0.4% excess mortality rate per cycle based on the 5% 6-month excess mortality used in NICE TA710 [17].

### Analysis of health economics trial data

The SETS aHUS trial collected quality of life data using the EQ-5D-5L questionnaire and resource use data from disease management using health care utilisation questionnaires at baseline and months 1, 3, 6, 9, 12, 18, and 24 of follow-up. Data were collected for the trial population withdrawing from eculizumab (n = 28) and a control cohort maintaining eculizumab during the trial follow-up (n = 11), (Figure 3).

**Figure 3.**
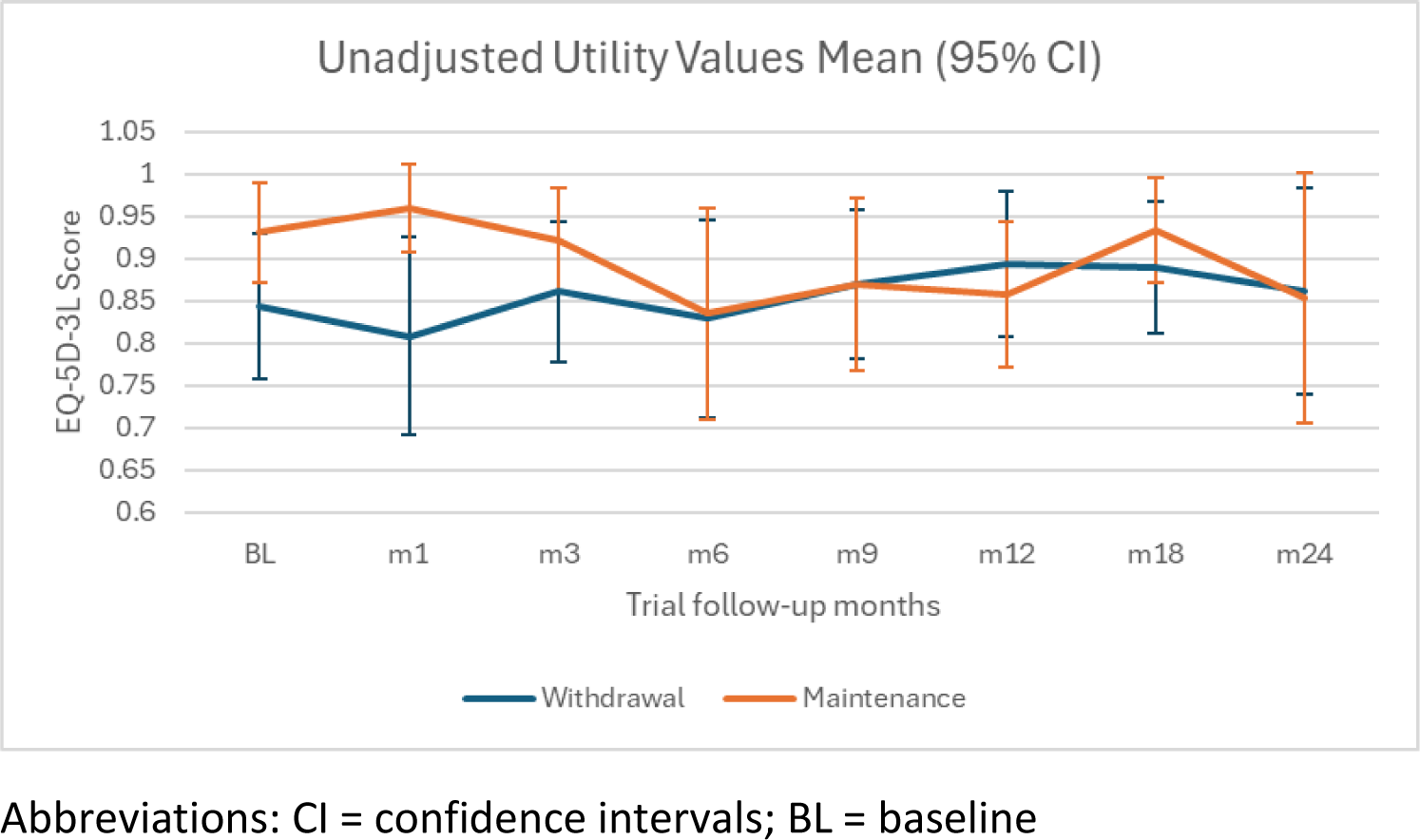
Unadjusted EQ-5D-3L scores by SETS aHUS follow-up: withdrawal versus maintenance group.

Missing data was a primary issue during data analysis, as a complete case analysis of total QALYs showed that 61% of participants had at least one missing EQ-5D-5L in the withdrawal group (11 complete records), and 91% of participants had at least one missing EQ-5D-5L in the control group (1 complete record). Similar levels of missingness were observed in the total healthcare use data with 57% missing at least one questionnaire in the withdrawal group (12 complete records), and 91% missing at least one questionnaire in the control group (1 complete records). Complete case estimates were presented descriptively as mean total QALYs and mean total costs for each treatment arm.

Due to the small sample size in the trial and the large proportion of missing data, available case analysis was deemed more appropriate to inform the utility changes for withdrawing from treatment and disease relapse [18]. A multivariate random effects model was used instead controlling for time at follow-up, baseline utility, age, and gender. Monthly health care resource use data were also assessed using available case analysis, with a random effects model controlling for time, baseline costs, age, and gender. Statistical analyses were performed in STATA (V18) and figures drawn using Microsoft Office Excel (V2016). The EQ-5D and resource use data of one patient were considered as outliers (see Figure 4 and Figure 5), since the small sample size makes the analysis vulnerable to outlier effects the scenario analysis includes the impact of controlling for the outlier on costs and QALYs.

**Figure 4.**
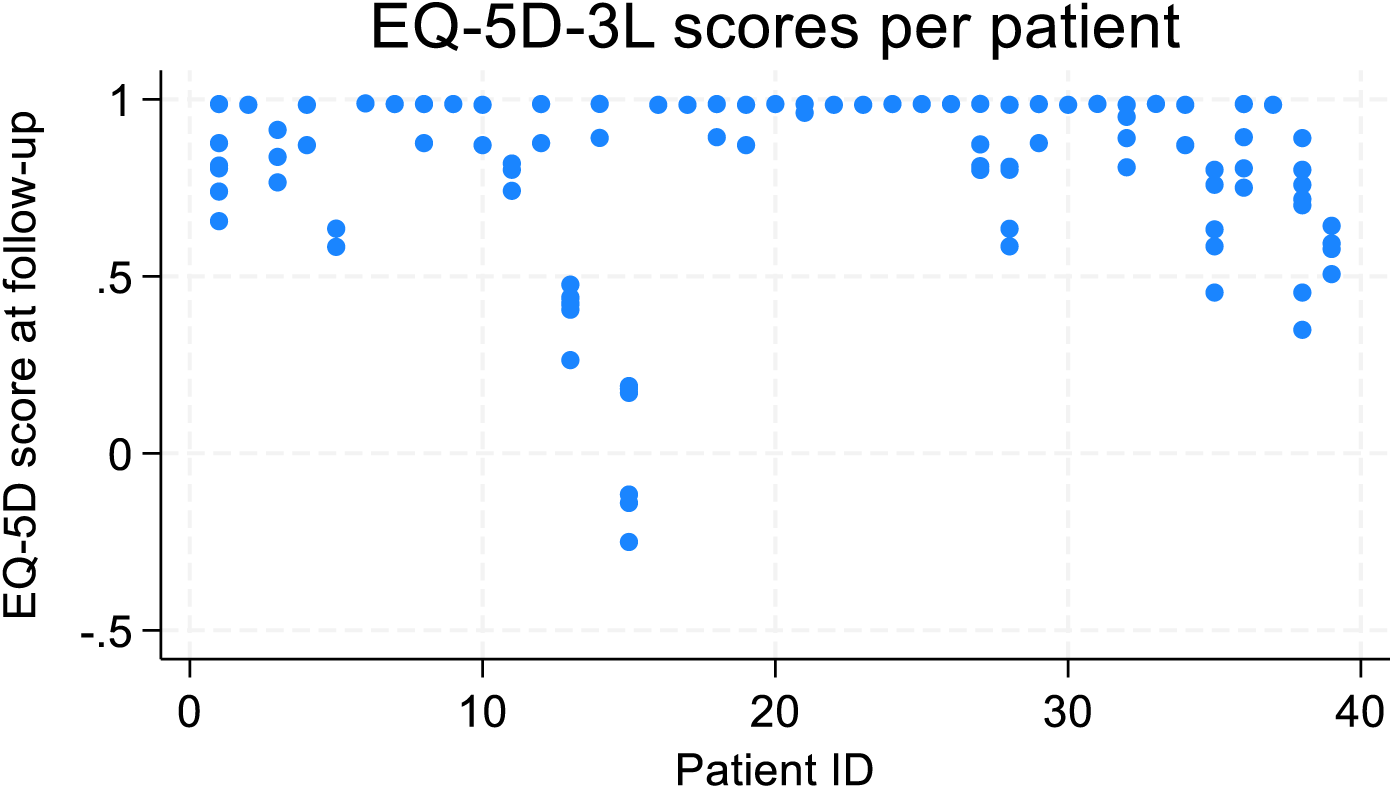
EQ-5D-3L scores per patient from the SETS aHUS trial.

**Figure 5.**
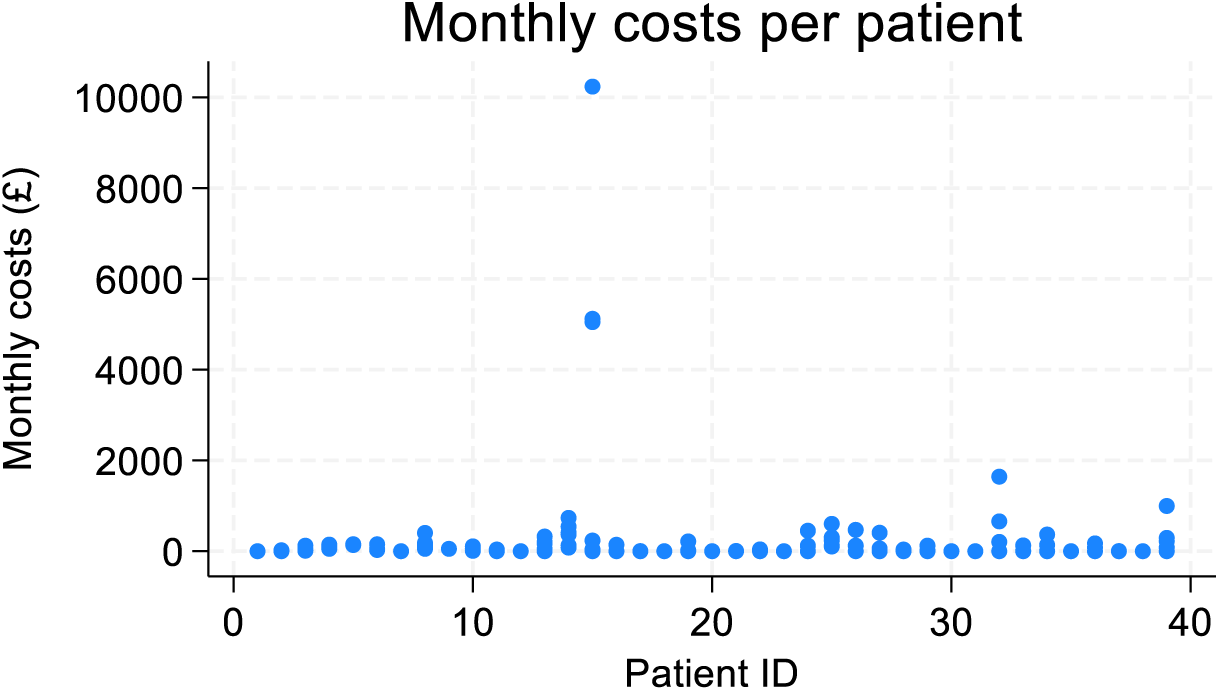
Monthly costs per patient from the SETS aHUS trial.

### Health outcomes

Health outcomes in the model were measured using quality adjusted life years (QALYs). Quality of life weights were calculated based on patient responses to the EQ-5D-5L questionnaire from the SETS aHUS trial [1, 19]. Responses were transformed into EQ-5D-3L scores using UK population values adjusted by age and gender [20, 21].

Data from the aHUS trial was used to inform quality of life in the CKD 0-2 and CKD 3 health states, and utility changes from treatment withdrawal and disease relapse. For more advanced disease stages not covered by the 2-year trial follow-up (CKD stage 4 or higher) quality of life values were sourced from the published literature and the technology assessments approved by NICE on aHUS [8, 9, 11, 12]. A scenario analysis assessed the impact of using quality of life weights from the literature rather than the trial values.

### Resource use and costs

All costs were presented using 2022 British pounds Sterling (£ GBP). Treatment costs with eculizumab and later ravulizumab were sourced from the BNF, including treatment initiation and treatment maintenance schedules [22]. Delivery costs beyond drug acquisition were sourced from the trial for the hospital setting, since delivery costs at the homecare setting are currently funded by Alexion Pharma UK [23].

The resource use of disease monitoring during both treatment maintenance and withdrawal were sourced from NICE TA710 and cost values were updated for inflation to 2022 values [24]. Disease management costs at CKD stage 4 and ESRD were also sourced from NICE TA710, while kidney transplant and post-transplantation costs were sourced from their respective NHS Tariffs, with additional health care staff costs sourced from Jones et al. 2023 [24–26].

Data from the SETS aHUS trial was used to inform disease management costs for CKD 0-2 and CKD 3 patients withdrawing from eculizumab, and for the control group maintaining treatment [1]. Cost data in the SETS aHUS trial health care utilisation questionnaires were used to inform additional health care costs from treatment maintenance, withdrawal, and disease relapse in the model [1].

### Perspective

Costs were included from an NHS and personal social services (PSS) perspective. Outcomes were self-reported by patients through questionnaires measuring changes in health-related quality of life. The economic analysis was carried using a lifetime horizon, with an annual discount rate on all outputs of 3.5% [27].

### Model-based analysis

Cost-utility analyses were conducted at the cohort level and presented in an incremental analysis over a patient’s lifetime. A deterministic sensitivity analysis was used to identify the parameters with the largest impact on cost-effectiveness, and a probabilistic analysis was conducted using the parametric distribution considered to best estimate the uncertainty around each input parameter [28] (e.g. the Gamma distribution primarily for cost parameters, the Beta distribution for probabilities and proportions, see Table 1 for more details). When estimates of variation were not available, a 10% standard error around the mean was assumed.

**Table 1.** Model parameters.

| Parameter | Mean or % | SE | PSA Distribution | Source |
| --- | --- | --- | --- | --- |
| Starting age (years) | 20 |  |  | Assumption |
| Males | 51% |  |  | [7] |
| **Disease distribution** |  |  |  |  |
| % of CKD 0-2 | 96% | 9.6% | Beta | [7] |
| % of CKD 3 | 4% |  |  | [7] |
| **Disease progression after relapse** |  |  |  |  |
| No treatment |  |  |  |  |
| CKD 0-2 to CKD 3 | 25% | 2.5% | Beta | [7] |
| CKD 3 to CKD 4 | 25% | 2.5% | Beta | [7] |
| On treatment |  |  |  |  |
| CKD 0-2 to CKD 3 | 0 |  |  | Assumption |
| CKD 3 to CKD 4 | 0 |  |  | Assumption |
| **Disease progression to advanced disease per cycle** |  |  |  |  |
| CKD 4 to ESRD | 2.55% | 0.25% | Beta | [24] |
| ESRD to kidney transplant | 3.25% | 0.32% | Beta | [2] |
| Transplant failure | 11.64% | 2.72% | Lognormal | [5] |
| **Excess mortality per cycle** |  |  |  |  |
| ESRD | 0.41% | 0.04% | Normal | [24] |
| Transplant | 0.41% | 0.04% | Normal | Assumption |
| Post-Transplant | 0.41% | 0.04% | Normal | Assumption |
| **Utility values** |  |  |  |  |
| Baseline CKD 0-3 | 0.95 | 0.07 | Beta | [7] |
| Transplant procedure | 0.66 | 0.07 | Beta | [2] |
| **Disutilities** |  |  |  |  |
| Treatment withdrawal | 0.02 | 0.03 | Normal | [7] |
| Disease relapse | -0.01 | 0.04 | Normal | [7] |
| CKD 4 | -0.15 | 0.02 | Normal | [24] |
| ESRD | -0.21 | 0.02 | Normal | [24] |
| Post-Transplant | -0.21 | 0.02 | Normal | [24] |
| **Drug acquisition costs** |  |  |  |  |
| Eculizumab | £3,150 |  |  | [22] |
| Ravulizumab | £16,621 |  |  | [22] |
| % Home delivery | 71% | 7.1% | Beta | SETS aHUS nurses [7] |
| Hospital drug delivery | £1,429 | 142.90 | Gamma | SETS aHUS nurses [7] |
| Antibiotic medications | £7 | 2.59 | Gamma | [29] |
| **Disease monitoring costs per cycle** |  |  |  |  |
| Maintenance | £102 | 10.24 | Gamma | [24, 26] |
| Withdrawal month 1 | £424 | 42.37 | Gamma | [24, 26] |
| Withdrawal month 2- 8 | £215 | 21.53 | Gamma | [24, 26] |
| Withdrawal month 8 onwards | £111 | 11.11 | Gamma | [24, 26] |
| **Disease management costs per cycle** |  |  |  |  |
| Treatment withdrawal | £19 | 96.77 | Gamma | [7, 25] |
| Disease relapse | £2 | 185.97 | Gamma | [7, 25] |
| CKD 4 | £18 | 1.77 | Gamma | [24] |
| ESRD | £24 | 2.36 | Gamma | [24] |
| Kidney transplant | £16 037 | 1 564.41 | Gamma | [24, 25] |
| Post-Transplant | £306 | 32.14 | Gamma | [24, 25] |
| Abbreviations: CKD = chronic kidney disease stage; ESRD = end stage renal disease; PSA = probabilistic sensitivity analysis; SE = standard deviation. |  |  |  |  |

The scenario analysis explored the impact on cost-effectiveness estimates from: an optimistic and pessimistic risk to restart treatment over time using the Gompertz and exponential distribution respectively; replacing the quality-of-life weights from the trial for values sourced from the literature; controlling for a patient identified as an extreme outlier in quality of life and resource use data from the trial; and reducing the price of eculizumab by 50%.

We also explored a scenario comparing lifelong treatment with an alternative to the current treatment withdrawal strategy, where patients who relapse receive a three-month course of eculizumab before stopping treatment again, based on some of the patterns observer by Wijnsma et al, (2018) [3]. In this scenario, patients can restart treatment as many times as they need rather than maintaining lifelong treatment after the first relapse. As this strategy was not assessed within the main SETS aHUS trial, clinical effectiveness in this scenario was based on the assumptions that the risk of a future relapse was the same as the risk of the first relapse, and that patients on treatment were at no risk of CKD stage deterioration.

The 2015 guidelines on the standard care of aHUS were updated in 2021 to include ravulizumab as an alternative C5 inhibitor in the treatment pathway [24]. As ravulizumab was not delivered during the SETS aHUS study, a scenario analysis explored a comparison of lifelong treatment versus treatment withdrawal and monitoring using ravulizumab instead of eculizumab to treat aHUS. This followed the assumption of clinical equivalence between eculizumab and ravulizumab in NICE TA 710 [11, 24].

Table 1 summarises the parameters used to inform the base-case economic model.

## Results

### Summary of main results

#### Trial-based quality of life and treatment effects

Average total QALYs were 1.63 (SE = 0.17) for the whole analysis sample during the 2-year trial duration of the SETS aHUS trial. For the trial population (withdrawal arm) total QALYs were 1.6 (SE = 0.18), and for the control arm 1.94 (no SE calculable). Results from the linear random effects model showed a positive but non-statistically significant difference in utility values for the withdrawal arm (0.02, p = 0.59), and a negative but non-statistically significant difference from disease relapse (−0.01, p = 0.84), (Table 2).

**Table 2.**
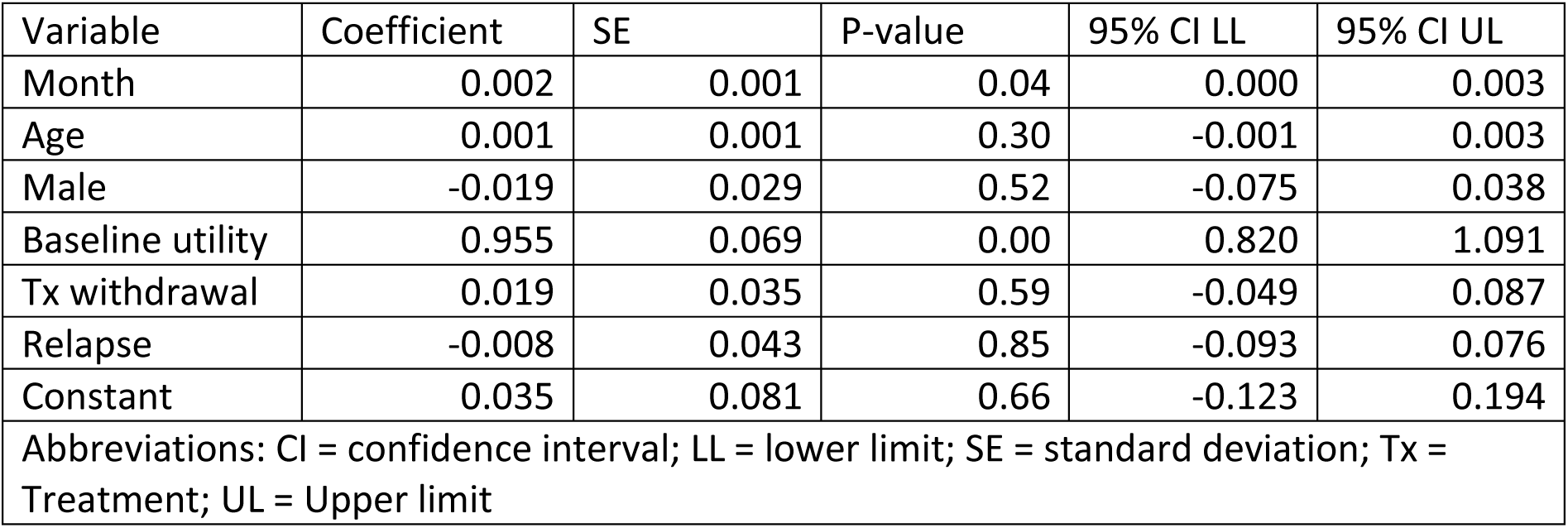
Random effects results for EQ-5D-3L utility scores - SETS aHUS trial.

| Variable | Coefficient | SE | P-value | 95% CI LL | 95% CI UL |
| --- | --- | --- | --- | --- | --- |
| Month | 0.002 | 0.001 | 0.04 | 0.000 | 0.003 |
| Age | 0.001 | 0.001 | 0.30 | -0.001 | 0.003 |
| Male | -0.019 | 0.029 | 0.52 | -0.075 | 0.038 |
| Baseline utility | 0.955 | 0.069 | 0.00 | 0.820 | 1.091 |
| Tx withdrawal | 0.019 | 0.035 | 0.59 | -0.049 | 0.087 |
| Relapse | -0.008 | 0.043 | 0.85 | -0.093 | 0.076 |
| Constant | 0.035 | 0.081 | 0.66 | -0.123 | 0.194 |
| Abbreviations: CI = confidence interval; LL = lower limit; SE = standard deviation; Tx = Treatment; UL = Upper limit |  |  |  |  |  |

#### Trial-based resource use

Average total costs were £2 012 (SE = 1 580) for the whole analysis sample during the 2-year trial duration of the SETS aHUS trial. For the trial population (withdrawal arm) total costs were £2 175 (SE = 1 709), and for the control arm £54 (no SE calculable). Results from the linear random effects model showed a positive but non-statistically significant difference in average monthly costs for the withdrawal arm (£41, p = 0.85), and a positive but non-statistically significant difference from disease relapse (£5, p = 0.99), (Table 3).

**Table 3.** Random effects results for monthly health care utilisation - SETS aHUS trial.

| Variable | Coefficient | SE | P-value | 95% CI LL | 95% CI UL |
| --- | --- | --- | --- | --- | --- |
| Month | 11.51 | 7.38 | 0.12 | -2.96 | 25.97 |
| Age | -2.38 | 4.94 | 0.63 | -12.07 | 7.31 |
| Male | 266.46 | 173.50 | 0.13 | -73.58 | 606.51 |
| Baseline utility | 0.19 | 0.18 | 0.28 | -0.16 | 0.55 |
| Tx withdrawal | 41.08 | 209.67 | 0.85 | -369.86 | 452.02 |
| Relapse | 5.21 | 402.93 | 0.99 | -784.51 | 794.93 |
| Constant | -377.71 | 326.94 | 0.25 | -1018.51 | 263.09 |
| Abbreviations: CI = confidence interval; LL = lower limit; SE = standard deviation; Tx = Treatment; UL = Upper limit |  |  |  |  |  |

### Cost-effectiveness results

Table 4 shows the deterministic and probabilistic cost-effectiveness results of the base-case analysis. Probabilistic results showed that stopping eculizumab with a disease monitoring strategy led to average cost-savings of £4 188 361 (incremental costs 95% CrI: -£6 390 713 to -£675 511) per patient and an increase in QALYs of 0.22 (incremental QALYs 95% CrI: −0.7 to 1.25 QALYs) compared with lifelong maintenance of eculizumab. Stopping eculizumab also led to a reduction in survival of 0.0005 LYs (incremental LYs 95% CrI: −0.003 to 0).

**Table 4.**
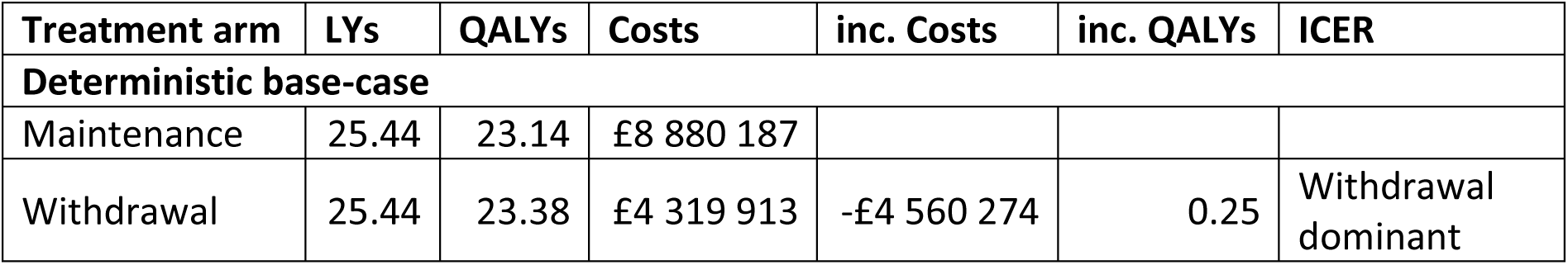
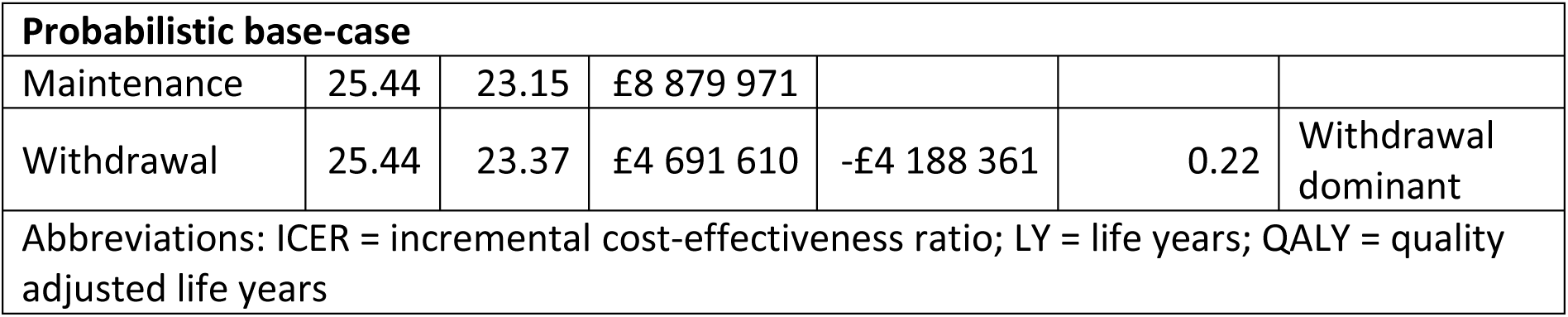
Base-case model results.

#### Effects of uncertainty

Results from the probabilistic sensitivity analysis showed that a treatment withdrawal strategy had a 100% probability of being cost-effective over all values for society’s willingness to pay for a QALY up to £250 000 and was more efficient and less costly in 71% of the simulations (Figure 6). A one-way sensitivity analysis on the deterministic incremental net monetary benefit (NMB) showed that the most impactful parameters on the cost-effectiveness results were the risk of treatment reinitiation after withdrawal, the initial proportion of CKD 0-2 patients versus more severe CKD stages at withdrawal, and the cost of eculizumab. The most impactful parameter for quality of life was the disutility of eculizumab withdrawal. The incremental NMB of eculizumab withdrawal was £4 567 661 for a £30 000 per QALY threshold. Results are presented in Figures 7, 8, and 9.

**Figure 6.**
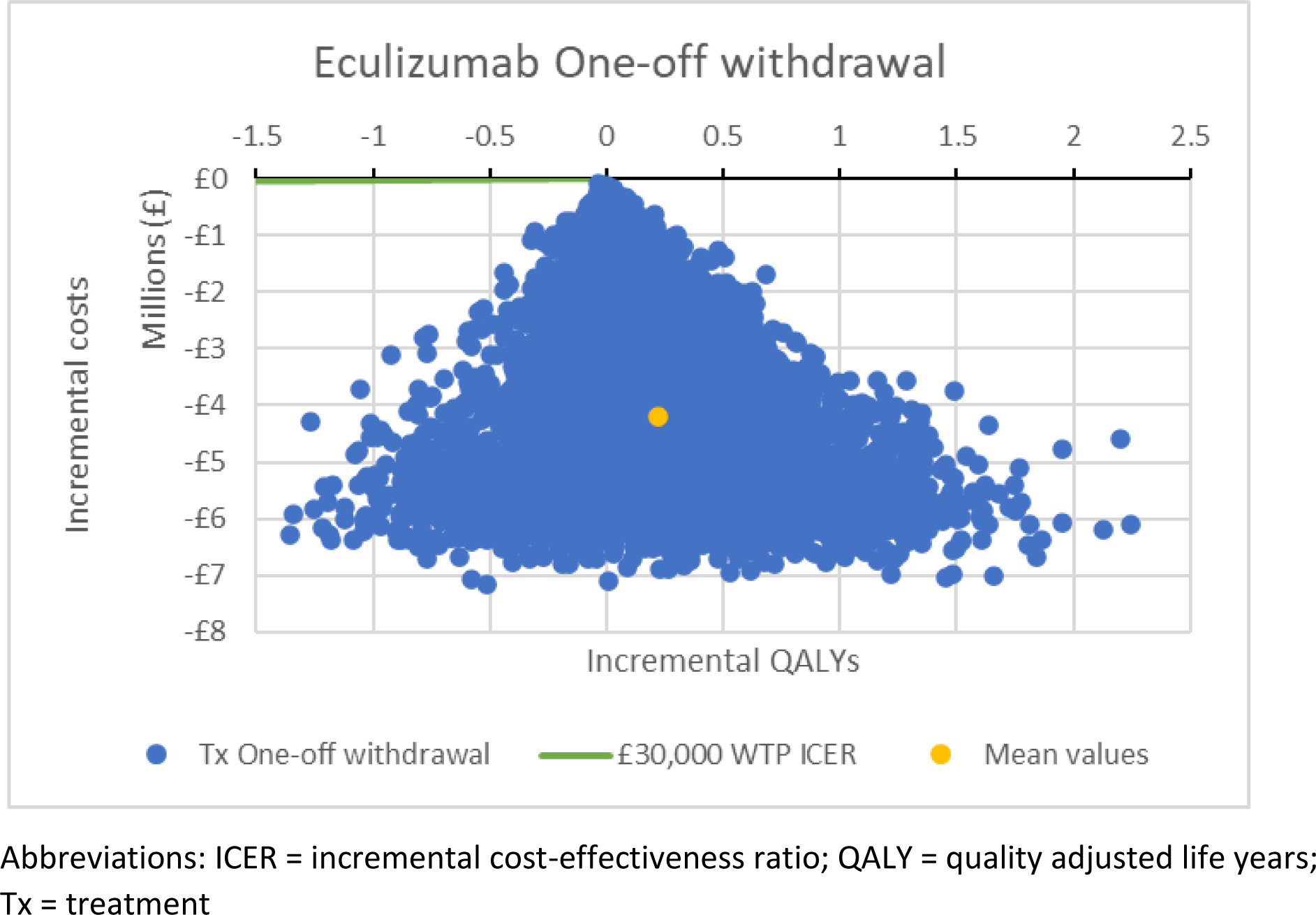
Cost-effectiveness plane.

**Figure 7.**
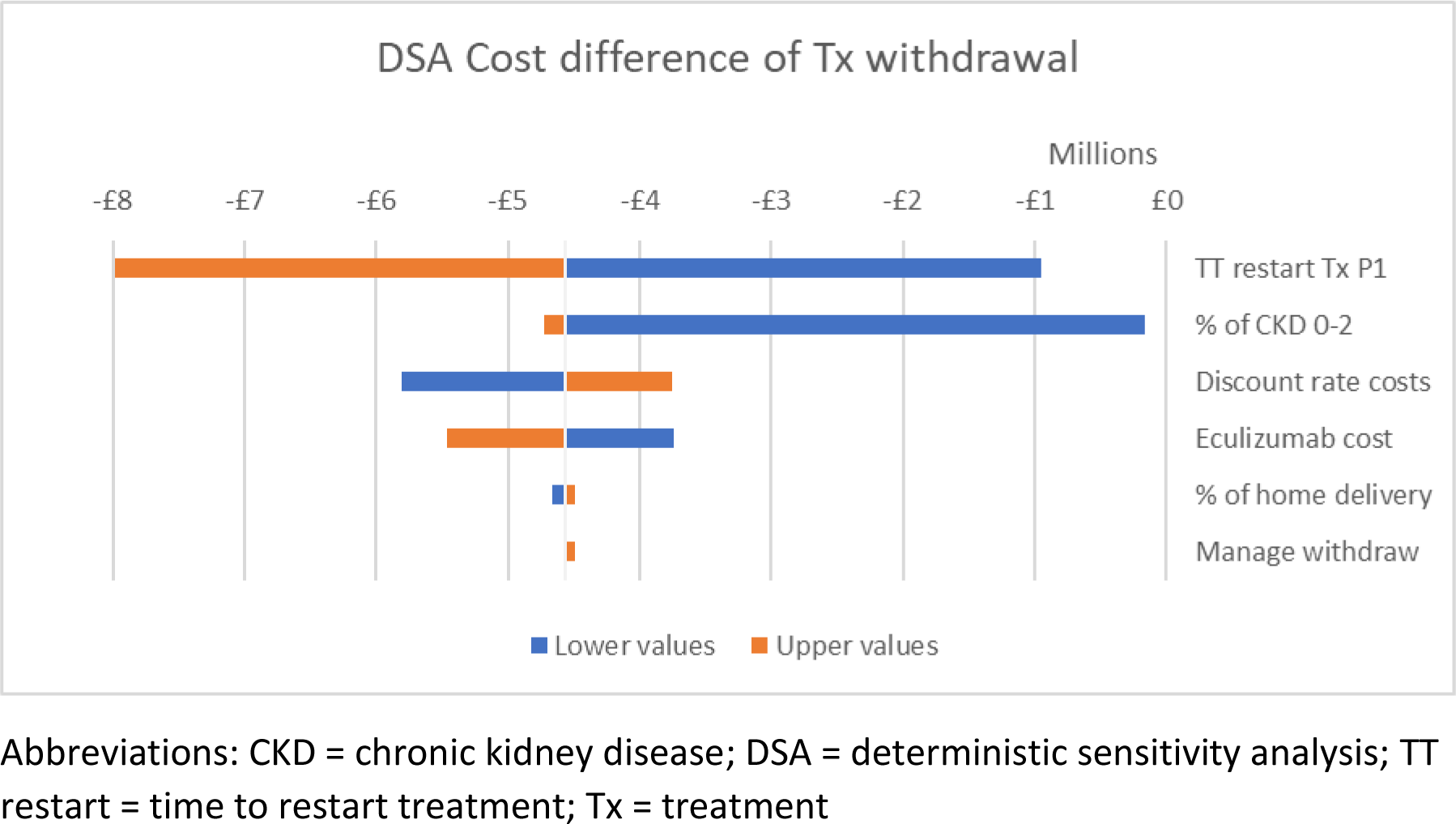
Deterministic sensitivity analysis on the cost difference of treatment withdrawal.

**Figure 8.**
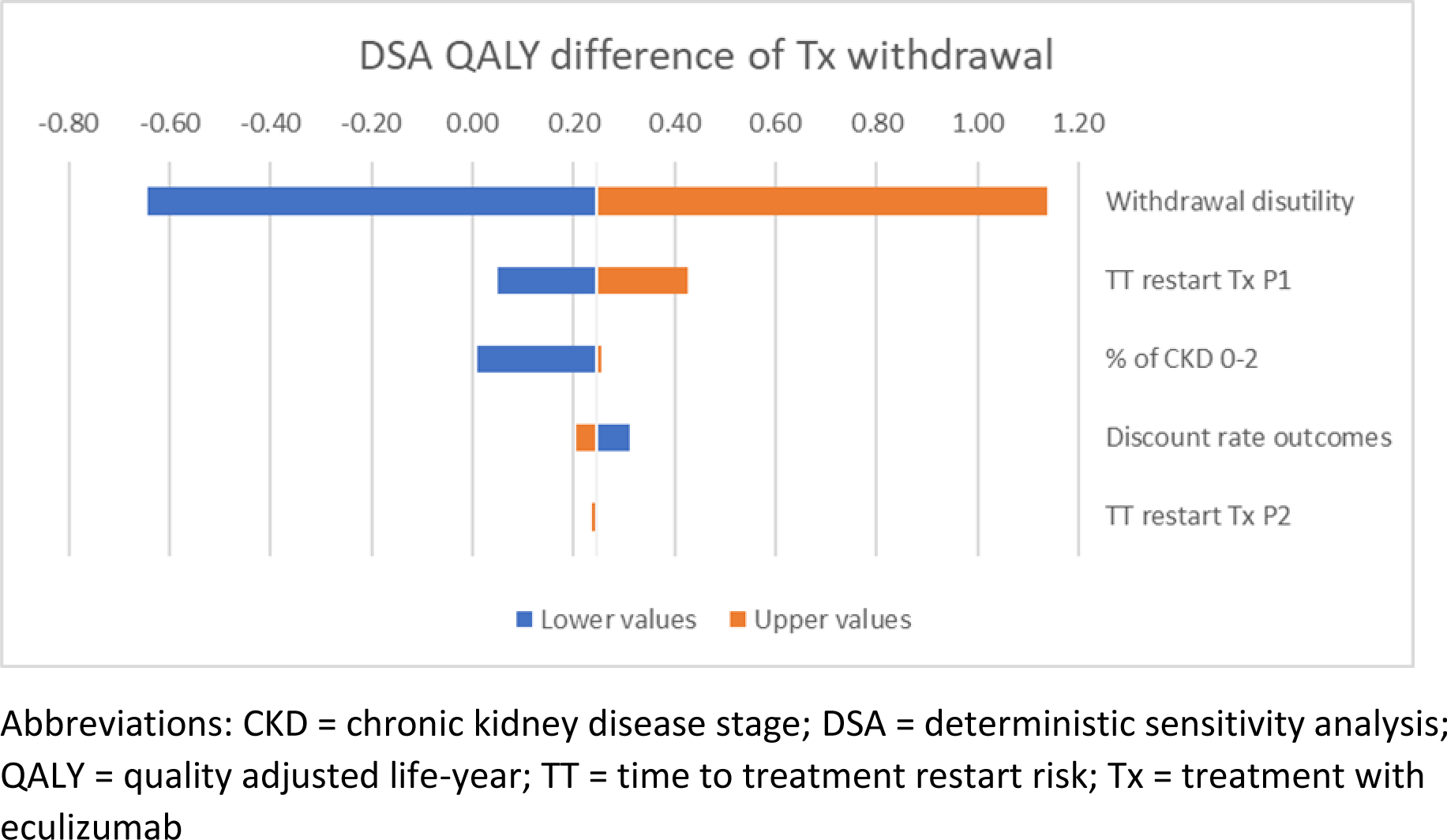
Deterministic sensitivity analysis on the QALY difference of treatment withdrawal.

**Figure 9.**
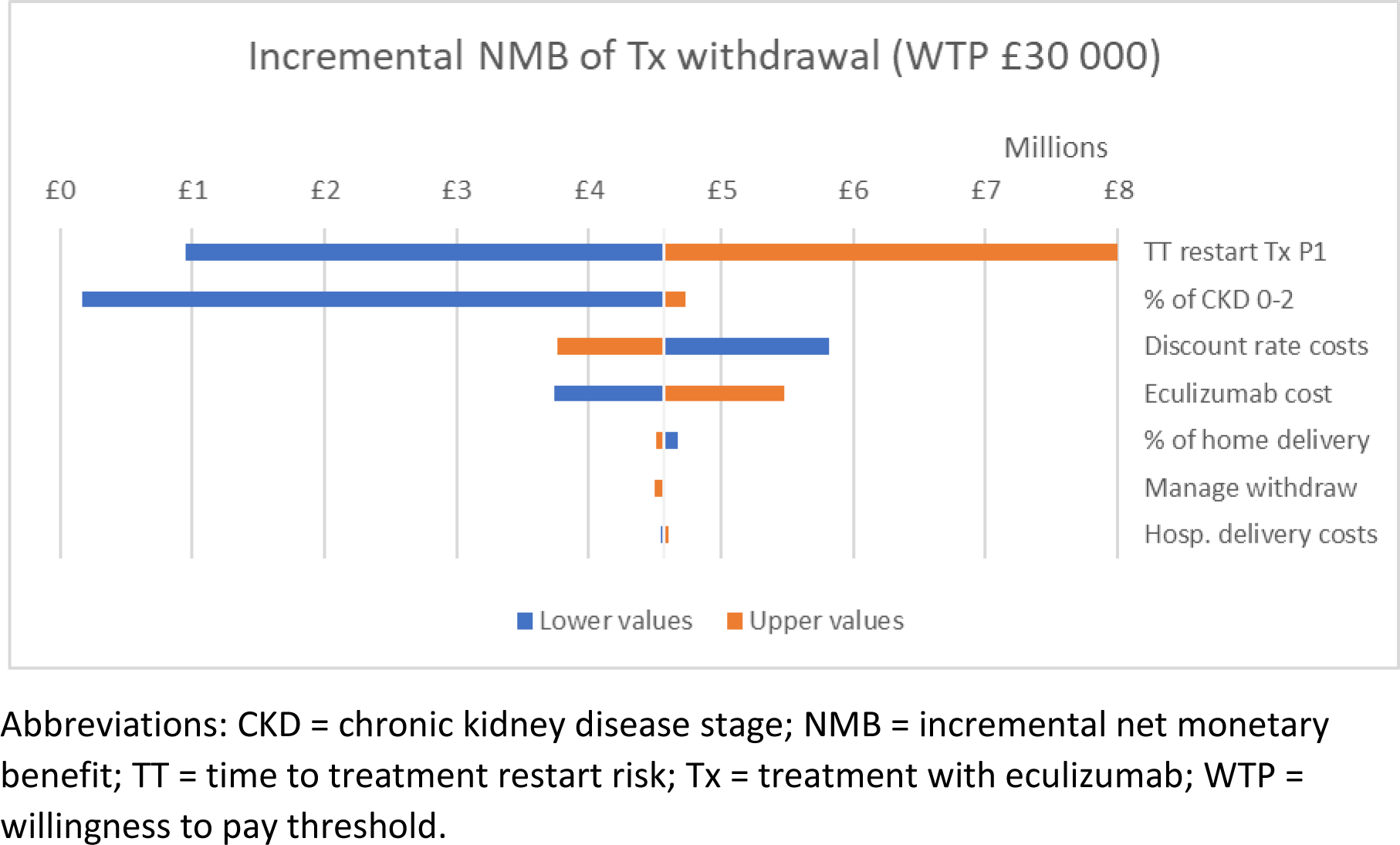
Deterministic sensitivity analysis for the incremental net monetary benefit of treatment withdrawal.

The scenario analysis showed that a strategy allowing for multiple withdrawals after reinitiation, the use of trial utility estimates after controlling for an outlier, and the risk or reinitiation over time have the biggest impact on effectiveness; while halving the price of eculizumab, the risk of reinitiation over time, and the multiple withdrawal strategy had the largest effect on costs (Table 5).

**Table 5.** Scenario analysis.

| Scenarios | LYs | QALYs | Costs | Inc. QALYs | Inc. Costs | ICER |
| --- | --- | --- | --- | --- | --- | --- |
| **Deterministic base-case** |  |  |  |  |  |  |
| Maintenance | 25.44 | 23.14 | £8 880 187 |  |  |  |
| Withdrawal | 25.44 | 23.38 | £4 319 913 | 0.25 | -£4 560 274 | Withdrawal dominant |
| **Multiple withdrawals** |  |  |  |  |  |  |
| Maintenance | 25.44 | 23.14 | £8 880 187 |  |  |  |
| Withdrawal | 25.44 | 23.60 | £ 350 379 | 0.46 | -£8 529 808 | Withdrawal dominant |
| **Gompertz distribution Tx restart (Favourable)** |  |  |  |  |  |  |
| Maintenance | 25.44 | 23.14 | £8 880 187 |  |  |  |
| Withdrawal | 25.44 | 23.52 | £1 732 504 | 0.38 | -£7 147 683 | Withdrawal dominant |
| **Exponential distribution Tx restart (Unfavourable)** |  |  |  |  |  |  |
| Maintenance | 25.44 | 23.14 | £8 880 187 |  |  |  |
| Withdrawal | 25.44 | 23.29 | £6 039 759 | 0.16 | -£2 840 428 | Withdrawal dominant |
| **Baseline QALYs from the literature** |  |  |  |  |  |  |
| Maintenance | 25.44 | 24.31 | £8 880 187 |  |  |  |
| Withdrawal | 25.44 | 24.56 | £4 319 913 | 0.25 | -£4 560 274 | Withdrawal dominant |
| **Baseline QALYs controlling for an extreme outlier** |  |  |  |  |  |  |
| Maintenance | 25.44 | 22.87 | £8 880 187 |  |  |  |
| Withdrawal | 25.44 | 23.01 | £4 319 913 | 0.14 | -£4 560 274 | Withdrawal dominant |
| **Direct costs controlling for an extreme outlier** |  |  |  |  |  |  |
| Maintenance | 25.44 | 23.14 | £8 880 187 |  |  |  |
| Withdrawal | 25.44 | 23.38 | £4 316 332 | 0.25 | -£4 563 855 | Withdrawal dominant |
| **Ravulizumab treatment** |  |  |  |  |  |  |
| Maintenance | 25.44 | 23.14 | £8 585 465 |  |  |  |
| Withdrawal | 25.44 | 23.38 | £4 185 779 | 0.25 | -£4 399 687 | Withdrawal dominant |
| **2-year time horizon** |  |  |  |  |  |  |
| Maintenance | 1.96 | 1.86 | £ 841 835 |  |  |  |
| Withdrawal | 1.96 | 1.90 | £ 237 410 | 0.03 | -£ 604 425 | Withdrawal dominant |
| **10-year time horizon** |  |  |  |  |  |  |
| Maintenance | 8.58 | 8.11 | £3 107 300 |  |  |  |
| Withdrawal | 8.58 | 8.23 | £ 936 896 | 0.12 | -£2 170 404 | Withdrawal dominant |
| **50% eculizumab price reduction** |  |  |  |  |  |  |
| Maintenance | 25.44 | 23.14 | £4 616 089 |  |  |  |
| Withdrawal | 25.44 | 23.38 | £2 267 997 | 0.25 | -£2 348 092 | Withdrawal dominant |
| Abbreviations: ICER = incremental cost-effectiveness ratio; LY = life years; QALY = quality adjusted life years |  |  |  |  |  |  |

## Discussion

The aim of this economic analysis was to assess the cost-effectiveness of using a treatment stoppage strategy on aHUS patients with CKD stage 0 to 3, as an alternative to a lifelong treatment eculizumab maintenance strategy. Data from the trial on patient quality of life, resource use, and clinical safety of eculizumab withdrawal were used to inform an economic model assessing the long-term cost-effectiveness impact for each patient.

The base-case analysis showed that a treatment withdrawal strategy with disease monitoring has the potential to improve patient quality of life on average by 0.22 QALYs and reduce lifetime costs by £4 188 361 compared with the lifetime maintenance of eculizumab. Model inputs included the risks of treatment reinitiation and disease progression after reinitiation, where the treatment withdrawal strategy led to a small average reduction in length of life of 0.0005 LYs (equivalent to 4 hours over a lifetime). The withdrawal and monitoring strategy was on average cost-saving, with a 71% probability of being more effective and less costly than treatment maintenance. The deterministic analysis identified the risk of relapse, the initial proportion of patients with CKD 0-2 versus CKD 3, and the cost of eculizumab as the primary drivers of cost-effectiveness.

Although previous models have attempted to include treatment withdrawal and restart [8, 11], our model is the first to use trial data to inform the time from withdrawal to restarting treatment using the SETS aHUS trial results [1]. This further showed that models relying on a constant hazard over time assumption for restarting treatment are potentially pessimistic, while using parametric models allowed us to relax this assumption.

Our analysis further explored scenarios that included changing the C5 inhibitor used to treat aHUS to ravulizumab, or changing the retreatment strategy to allow further withdrawals. Moreover, although the withdrawal and disease monitoring strategy remained cost-effective in comparison to lifelong treatment, due to the scarcity of data available these analyses relied on strong assumptions. These results therefore indicate a potential for further research, as we could expect the cost of such research to be more than offset by cost savings in the use eculizumab or ravulizumab.

The primary limitations of the economic analysis stemmed from the small sample size in the SETS aHUS trial (given that aHUS is a rare disease), compounded by large proportions of missing data and the effects of extreme outliers. Moreover, the follow-up time of the SETS aHUS trial (approximately 2 years after stopping treatment) was significantly shorter compared to the scope of the model (lifetime of a 20-year-old patient), nevertheless, our analysis suggested likely gains in quality of life and evidence of cost-savings over the first 2 and 10 years of treatment withdrawal. Our analyses further incorporated the imprecision in estimates caused by these uncertainties, and it is reassuring that our sensitivity analyses confirmed the conclusions drawn from the study, moreover there is potential for the use of real-world evidence to further strengthen the evidence available.

### Conclusions

The long-term benefits of lifelong treatment with eculizumab are still uncertain. Results from our model-based cost-effectiveness analysis suggest that eculizumab withdrawal coupled with disease monitoring for eligible aHUS patients will result in considerable cost savings and a potential improvement in patient quality of life, with very little impact on survival. Consequently, treatment withdrawal is highly likely to be cost-effective. This conclusion remained robust to the wide range of scenarios considered.

## Data Availability

All data produced in the present study are available upon reasonable request to the authors

## Data availability statement

## Acknowledgements

We acknowledge the help of Mr Eoin Moloney and Dr Paul Tappenden for assisting with model conceptualisation.

This report presents independent research commissioned by the National Institute for Health Research (NIHR). The views and opinions expressed by authors in this publication are those of the authors and do not necessarily reflect those of the NHS, NIHR, the Medical Research Council, the NIHR Central Commissioning Facility, the NIHR Evaluation, Trials and Studies Coordinating Centre, the Health Technology Assessment programme or the Department of Health.

## Authors’ contributions

NS and YO conceptualized and designed the study. GO was responsible for data analysis and interpretation of the results. All authors contributed to the interpretation of the results, preparation and review of the manuscript, and the approval of the final manuscript for publication.

